# Cardiac remodeling in moderate aortic stenosis

**DOI:** 10.1101/2024.02.19.24303060

**Authors:** Marilena Giannoudi, Henry Procter, Sindhoora Kotha, Nicholas Jex, Amrit Chowdhary, Sharmaine Thirunavukarasu, Peter Swoboda, Sven Plein, Richard M. Cubbon, Hui Xue, Ladislav Valkovič, Peter Kellman, Marc R. Dweck, John P Greenwood, Eylem Levelt

**Affiliations:** University of Leeds, Biomedical Imaging Science Department, Leeds Institute of Cardiovascular and Metabolic Medicine, LS2 9JT, United Kingdom; National Heart, Lung, and Blood Institute, National Institutes of Health, DHHS, 10 Center Drive MSC-1061, Bethesda, MD, 20892, USA; Centre for Clinical Magnetic Resonance Research (OCMR), RDM Cardiovascular Medicine, University of Oxford, Oxford, United Kingdom; Department of Imaging Methods, Institute of Measurement Science, Slovak Academy of Sciences, Bratislava, Slovakia; University of Edinburgh/BHF Centre for Cardiovascular Science, Chancellor’s Building, 49 Little France Crescent, Edinburgh, EH16 SU4, UK; Baker Heart and Diabetes Institute & Monash University, Melbourne, Australia

**Keywords:** Aortic stenosis, Moderate aortic stenosis, Cardiovascular magnetic resonance imaging, Myocardial remodeling, Myocardial perfusion

## Abstract

**Background:** Aortic stenosis (AS) accounts for substantial global morbidity and premature mortality even in moderate AS (Mod-AS). The mechanisms for this adverse prognosis in Mod-AS, however, remain poorly understood, although the myocardial remodeling response is thought to be critical. We aimed to prospectively assess myocardial remodeling, perfusion and energetics differences in Mod-AS and severe AS (Severe-AS).

**Methods:** Fifty-two Severe-AS and 25 Mod-AS patients and 18 demographically-matched controls underwent cardiovascular magnetic resonance and phosphorus-magnetic resonance spectroscopy to define left ventricular (LV) mass and function, global longitudinal shortening (GLS), rest and adenosine-stress myocardial blood flow (MBF), myocardial perfusion reserve (MPR), layer-specific perfusion metrics (subendocardial [Endo], subepicardial [Epi] MBF and MPR, and Endo-to Epi-MBF ratio [Endo/Epi]), myocardial scar on late gadolinium enhancement (LGE) imaging, and myocardial energetics (phosphocreatine:ATP ratio [PCr/ATP]).

**Results:** Compared to controls, from Mod-AS to Severe-AS there was a progressive increase in LV concentricity (LV-mass/LV-end-diastolic-volume)(controls:0.58[0.54,0.62], Mod-AS:0.74[0.64,0.84], Severe-AS:0.89[0.83,0.95]g/mL; *P*<0.0001), LV mass-index (controls: 46[40,51], Mod-AS: 58[51,65], Severe-AS: 70[65,75]g/m^2^; *P*<0.0001) and stepwise decline in GLS (controls:19.9[17.6,22.2], Mod-AS:17.7[16.6,18.8], Severe-AS:13.4[12.5,14.4]%; *P*<0.0001) with significant differences between Mod-AS and Severe-AS for all three comparisons.

Both stress MBF (controls:2.1[1.9,2.3], Mod-AS:1.9[1.6,2.2], Severe-AS:1.3[1.2,1.5]ml/min/g; *P*<0.0001) and MPR (controls:3.3[2.8,3.6], Mod-AS:2.8[2.4,3.2], Severe-AS:1.9[1.8,2.1]; *P*<0.0001) were only significantly reduced in Severe-AS compared to controls, with significant differences also detected between Mod-AS and Severe-AS. However, stress-endo-MBF (controls:2.0[1.8,2.3], Mod-AS:1.7[1.5,2.0], Severe-AS:1.2[1.1,1.3] ml/min/g; *P*<0.0001), stress-Endo/Epi (controls:1.00[0.93,1.07], Mod-AS:0.87[0.80,0.94], Severe-AS:0.81[0.75,0.82]; *P<*0.0001), rest-Endo/Epi (controls:1.12[1.10,1.14], Mod-AS:1.06[1.03,1.09], Severe-AS:1.03[1.02,1.06]; *P*<0.0001) and endo-MPR (controls:3.2[2.7,3.6], Mod-AS:2.5[2.1,2.9], Severe-AS:1.7[1.5,1.8]; *P<*0.0001) were all significantly reduced in both Mod-AS and Severe-AS.

Compared to controls, both AS groups showed significantly lower PCr/ATP (controls:2.2[2.0,2.3], Mod-AS:1.8[1.6,2.0], Severe-AS:1.7[1.6,1.8]; *P*<0.0001) and shorter 6-minute-walk-distance (controls:525[495,555], Mod-AS:420[375,465]m, Severe-AS:345[248,420]m; *P<*0.0001).

Only the Severe-AS group had evidence of non-ischemic myocardial scarring on LGE (2.9[0.0,6.2]%), which was detected in 65% (n=34) of patients. Neither group had evidence of ischemic scar.

The AS severity (peak aortic valve velocity) correlated with the stress-MBF (r=-0.45, *P*=0.0003), MPR (r=-0.44, *P*=0.0005) and GLS (r=-0.47, *P*=0.0001).

**Conclusions:** Moderate and severe AS are both associated with cardiac concentric hypertrophy, reductions in myocardial energetics, subendocardial hypoperfusion, and limitations in exercise distance. Patients with Severe-AS exhibit a more pronounced phenotype with worse LV hypertrophy, contractile dysfunction and myocardial scarring compared to patients with Mod-AS.

**CLINICAL PERSPECTIVES:** What is new:

- Patients with moderate aortic stenosis show cardiac concentric hypertrophy, reduction in myocardial energetics, shorter 6-minute walk distance compared to age-and sex-matched controls, but no evidence of myocardial scarring.
- Global myocardial perfusion metrics of rest and stress myocardial blood flow or myocardial perfusion reserve do not show significant reductions in patients with moderate aortic stenosis.
- The perfusion dynamics of the epicardial and endocardial layers differ in the presence of moderate aortic stenosis, with significant reductions in endocardial stress myocardial blood flow, rest and stress endocardial to epicardial myocardial blood flow ratio, and endocardial as well as epicardial myocardial perfusion reserve compared to controls.

**What Are the Clinical Implications?:**

- While milder compared to that seen in severe aortic stenosis, the degree of adverse myocardial remodeling and subendocardial hypoperfusion associated with moderate aortic stenosis may be clinically important for the cardiovascular clinical outcomes in patients with moderate aortic stenosis.
- Larger prospective serial studies and randomized trials are needed to better understand the mechanisms of high cardiovascular and all-cause mortality rates in patients with moderate aortic stenosis.

## INTRODUCTION

Aortic stenosis (AS) is the most common valvular heart disease in the Western world^1^. AS represents a progressive degeneration process which can have a decade-long asymptomatic subclinical stage^1^. The degree of valve restriction varies from mild, to moderate, to eventually severe with gradual worsening over time, in the absence of effective medical therapy capable of slowing disease progression. AS accounts for substantial global morbidity and premature mortality even in patients with less than severe AS^2,3^. Epidemiological studies suggest reduced survival rates in patients with moderate AS with 5-year mortality rates as high as 56%, which is nearly equivalent to severe AS^4,5^. Compared with no AS, the adjusted risk of all-cause mortality was shown to be 1.92-fold higher in moderate AS, and 2.27-fold higher in severe AS^6^. However, the mechanisms for this adverse prognosis in moderate AS remain poorly understood.

Emerging evidence suggests that the myocardial remodeling response to AS is critical for the long-term prognosis, with the extent of cardiac damage shown to be linked to worse prognosis, independent of valvular obstruction severity^7,8^. During the asymptomatic period, the heart compensates for the pressure overload with increased wall thickness, which maintains normal wall stress and contraction^9^. But with a long period of excessive left ventricular (LV) pressure loading, eventually the compensatory mechanisms fail, resulting in myocardial damage and symptoms of angina, syncope or heart failure.^9^ In this situation the only effective treatment is aortic valve replacement (AVR), either surgically (SAVR) or using a transcatheter approach (TAVR). The current guidelines recommend AVR in patients with severe AS in the presence of symptoms or evidence of cardiac dysfunction (LV ejection fraction <50% on cardiac imaging)^1,10^.

Cardiovascular magnetic resonance imaging (CMR) is the reference standard for the assessment of the myocardial remodeling observed in AS, including LV volumes, mass and wall thickness as well as changes in systolic function^7^. CMR also provides assessment of myocardial fibrosis using late gadolinium enhancement and T1 mapping techniques, which provide powerful prognostic information in patients with AS undergoing AVR^11^. CMR perfusion mapping allows pixel-wise quantification of global perfusion indices at rest and pharmacological stress including myocardial blood flow (MBF) in ml/g/min and myocardial perfusion reserve^12^. The perfusion dynamics of the epicardial and endocardial layers differ in severe AS, with preferential flow shifting from the endocardium to epicardium^13^. Automated in-line myocardial perfusion quantification using CMR also provides a comprehensive assessment of the endocardium and epicardium perfusion^14^. Moreover, ^31^phosphorus magnetic resonance spectroscopy (^31^P-MRS) allows non-invasive assessment of the myocardial energetic state^15^. Using ^31^P-MRS, multiple studies have shown myocardial energetic compromise as reflected by reductions in myocardial energetics index phosphocreatine to ATP (PCr/ATP) to be an important feature of the metabolic phenotype of the AS.

To date only few studies, and of small sample size, assessed and compared myocardial remodeling in patients with moderate and severe AS^16^. This prospective study aimed to test the hypothesis that not only patients with severe AS but also patients with moderate AS exhibit significant myocardial remodeling which may underpin their adverse prognosis compared to the general population^17,18^. Therefore, using CMR and ^31^P-MRS, cardiac phenotype differences between patients with moderate AS and severe AS in comparison to demographically matched volunteers with normal aortic valve function were assessed prospectively. Imaging assessments were supported by 6-minute walk tests and plasma N-terminal pro-B-type natriuretic peptide (NT-proBNP) levels.

## METHODS

### Study design and oversight

This single-center, prospective case-control study complied with the Declaration of Helsinki and was approved by National Research Ethics Committees (ref:18/YH/0168 for the severe AS cohort and ref:18/YH/0168 for the moderate AS cohort and healthy volunteers). The study was funded by the Wellcome Trust (Grant 207726/Z/17/Z). We have previously reported changes in myocardial energetics and perfusion parameters and LV structure and function in patients with severe AS (30 with and 65 without T2D) before and after AVR, and reported myocardial recovery differences after AVR between severe AS patients with and without T2D.

### Inclusion criteria

Adult patients with severe AS who had been referred for surgical or transcatheter AVR and patients with moderate AS who were under the routine clinical care of the local valve clinic at the Leeds Teaching Hospitals NHS Trust were eligible for inclusion. The diagnosis of severe AS was based on peak aortic forward flow velocity of greater than 4m/s on valve clinic echocardiography according to current cardiovascular society guidelines^1,10^. The diagnosis of moderate AS was based on peak aortic velocity of 3.0–3.9m/s or mean gradient of 20– 39mmHg and aortic valve area (AVA) >1 and <1.5cm^2^ on valve clinic echocardiography^28^. Controls were contemporary participants and were recruited from adverts posted at leisure activity walking groups in Yorkshire, United Kingdom.

The local patient population with moderate AS under regular valve clinic follow up consisted of 244 patients. Electronic health records were prescreened against the eligibility criteria and potentially eligible patients with moderate AS were invited for participation.

### Exclusion criteria

Participants were excluded if they had previous myocardial infarction, coronary artery bypass grafting or angioplasty, flow-limiting coronary artery disease (CAD), chronic obstructive lung disease, more than mild bystander valve disease, significant kidney dysfunction (estimated glomerular filtration rate [eGFR] <30mL/min/1.73m^2^), known heart failure or reduced left ventricular ejection fraction (<55%), cardiomyopathy (based on infiltrative diseases [e.g., amyloidosis], accumulation diseases [e.g., haemochromatosis, Fabry disease], or hypertrophic cardiomyopathy), or any contraindication to CMR scanning. As the research protocol included a 6-minute walk test, potential participants with mechanical or permanent mobility issues were excluded. All patients with severe AS were listed for AVR and had flow-limiting CAD excluded by invasive angiography. Prior myocardial infarction was excluded by late gadolinium enhancement imaging in all participants.

### Anthropometric measurements

Height and weight were recorded, and body mass index (BMI) was calculated. The blood pressure was recorded whilst seated over 10 minutes. A 12-lead electrocardiogram was recorded. A fasting blood sample was taken for assessments of full blood count, eGFR, lipid profile, glycated hemoglobin (HbA1c), renal function, lipid profile, glucose level, and NT-proBNP levels.

### Six-minute walk test

Functional exercise capacity was assessed using a 6-minute walk test according to established guidelines^19^. Participants were instructed to walk along a 30-meter corridor and cover the maximum achievable distance in 6 minutes under the supervision of investigators trained in conducting the test. At the end of 6 minutes, participants were asked to stop, and the distance walked was measured in meters.

### ^31^Phosphorus magnetic resonance spectroscopy

Magnetic resonance imaging (MRI) and ^31^P-MRS were performed on a 3.0 Tesla MR system (Prisma, Siemens, Erlangen, Germany). ^31^P-MRS was performed to assess myocardial PCr/ATP from a mid-ventricular septum voxel selected from a 3D MRS imaging matrix, with patients lying supine and a ^31^P transmitter/receiver cardiac coil (Rapid Biomedical GmbH, Rimpar, Germany) placed over the heart as previously described^20^. ^31^P-MRS data were acquired with a non-gated 3D acquisition-weighted chemical shift imaging (CSI) sequence.

### Cardiovascular magnetic resonance

The CMR protocol (Figure-1) consisted of cine imaging using a steady-state free precession sequence, native pre-and post-contrast T1 mapping, stress and rest perfusion and late gadolinium enhancement imaging.

**Figure 1:**
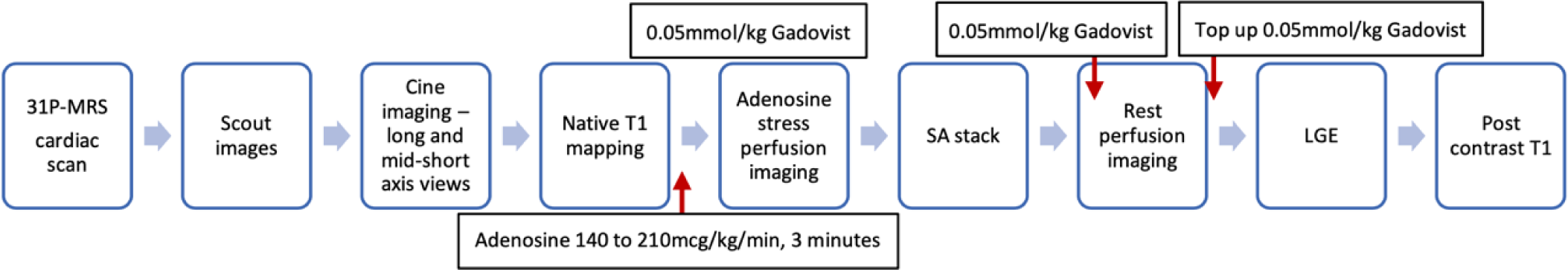
Multiparametric scan protocol. Cardiac ^31^P-MRS was followed by CMR, which included cine imaging, native pre-contrast, and native post-contrast T1 mapping, adenosine stress perfusion imaging and late gadolinium enhancement imaging.

Native T1 maps were acquired in 3 short-axis slices, using a breath-held modified look-locker inversion recovery acquisition as previously described^21^. Post-contrast T1 mapping was performed using the same approach 15 minutes after the last contrast injection.

Perfusion imaging used free-breathing, motion-corrected automated in-line perfusion mapping. Adenosine was infused at a rate of 140µg/kg/min, for a minimum of 3 minutes according to hemodynamic and symptomatic response as previously described^21^. Two trained cardiologists with advanced life support training monitored the patients during adenosine stress imaging, however adenosine stress was tolerated well by all patients during the stress perfusion studies. Late gadolinium enhancement was performed using a phase-sensitive inversion recovery sequence >8 minutes after contrast administration.

### Quantitate analysis of ^31^P-MRS and CMR data

All ^31^P-MRS and CMR post-processing analyses (with the exception of perfusion mapping and global longitudinal shortening [GLS] data) were performed offline. Perfusion mapping used artificial intelligence (Gadgetron Framework) for instant quantification of perfusion indices in addition to rest and stress MBF and MPR. Segmental layer-specific perfusion metrics (subendocardial [Endo], subepicardial [Epi] MBF and MPR, and Endo to Epi MBF ratio [Endo/Epi]) values were automatically obtained as an average of all pixel values of each of the 16 segments^22^. The 16 segments were further sub-divided transmurally to create endocardial and epicardial segments with corresponding rest and adenosine-stress MBF values and ratios to reflect transmural gradient. Global endocardial:epicardial (Endo/Epi) ratio was calculated by averaging the ratio across the 3 slices.

The ^31^P-MRS was performed offline by MG using software within the MATLAB version R2021a (MathWorks) as previously described.^20^

Cvi42 software (Circle cardiovascular imaging) was used to perform CMR analysis by MG and subsequently reviewed by EL. The images for biventricular and atrial volumes, biventricular and atrial function, as well LV maximal wall thickness were analyzed as previously described.^23^

Automated software provided GLS and mitral annular plane systolic excursion (MAPSE) measurements. T1 maps and extracellular volume (ECV) were analyzed using cvi42 software as previously described.^20^

After an initial visual assessment of the LV short axis stack images for presence of late gadolinium hyperenhancement, quantification was formally assessed using the tissue characterization tool within cvi42, as previously described.^24^ Late gadolinium hyperenhancement was defined as areas of signal intensity ≥5 SDs from normal myocardium and was expressed as the percentage of LV mass, quantified in a blinded fashion.

### Sample size

A priori sample size calculations were performed based on pilot data (LV concentricity index: relative ratio of LV mass to LV end-diastolic volume [EDV] and vasodilator-stress MBF) obtained from patients with moderate AS data (n=5) and healthy controls (n=15). These pilot study assessments showed mean LV mass to LVEDV ratio of 0.86±0.19 in patients with moderate AS versus 0.70±0.20 in controls. Based on these pilot data for the two groups, a minimum of 18 patients were needed to be recruited per group to detect a significant difference in LV mass to LVEDV ratio between patients with moderate AS and controls with 80% power at a 5% significance level on a two-sample t-test (calculations performed on ClinCalc.com software). A second sample size calculation was performed for comparisons of vasodilator-stress MBF between the two groups (Moderate-AS:1.74±0.24 versus controls:2.11±0.25ml/min/g). Based on these pilot data for the two groups, a minimum of 18 patients were needed to be recruited (9 per group) to detect a significant difference in vasodilator-stress MBF between patients with moderate AS with and controls with 90% power at a 5% significance level on a two-sample t-test (calculations performed on ClinCalc.com software). These targets were achieved in this study. In addition to these pre-specified comparisons other exploratory analyses were performed with due allowance for their exploratory nature. Power calculations were not performed for the exploratory endpoints.

### Statistical analysis

Statistical analysis was performed using GraphPad Prism software (version 10.0.3). All data were checked for normality using the D’Agostino-Pearson Test. Data are presented as mean ±95% confidence intervals (when normally distributed) or means with corresponding IQR (if nonparametric continuous variables). Categorical data are presented as numbers and percentages and compared with the Pearson χ^2^ test. All comparisons between >2 groups were performed by 1-way ANOVA with post hoc Bonferroni corrections. Differences in nonparametric variables were assessed using a Kruskal-Wallis test. Student t-test was used for comparison of normally distributed datasets and Mann-Whitney U test was used for non-parametric tests where data were obtained for only two groups. Bivariate correlations were performed using Pearson’s or Spearman’s method, as appropriate. A 2-sided *P* value of <0.05 was applied as indicating threshold of significance.

## RESULTS

### Participant demographics and clinical characteristics

Fifty-two patients with severe symptomatic AS awaiting AVR (18 with T2D), 25 patients with moderate AS (8 with T2D) under routine clinical monitoring and 18 controls with no valvular dysfunction or any other cardiovascular disease (5 with T2D) were prospectively recruited between September 2022 and October 2023 (supplementary materials figure, CONSORT diagram) were recruited. There were no significant differences in sex distributions or T2D prevalence across all study groups. The moderate AS (Mod-AS) and control groups were matched in age; however, the severe AS group (Severe-AS) were older (Table-1). Patients with severe AS were more symptomatic with 90% of the group classified NYHA Class II to IV (Table-1).

**Table 1:**
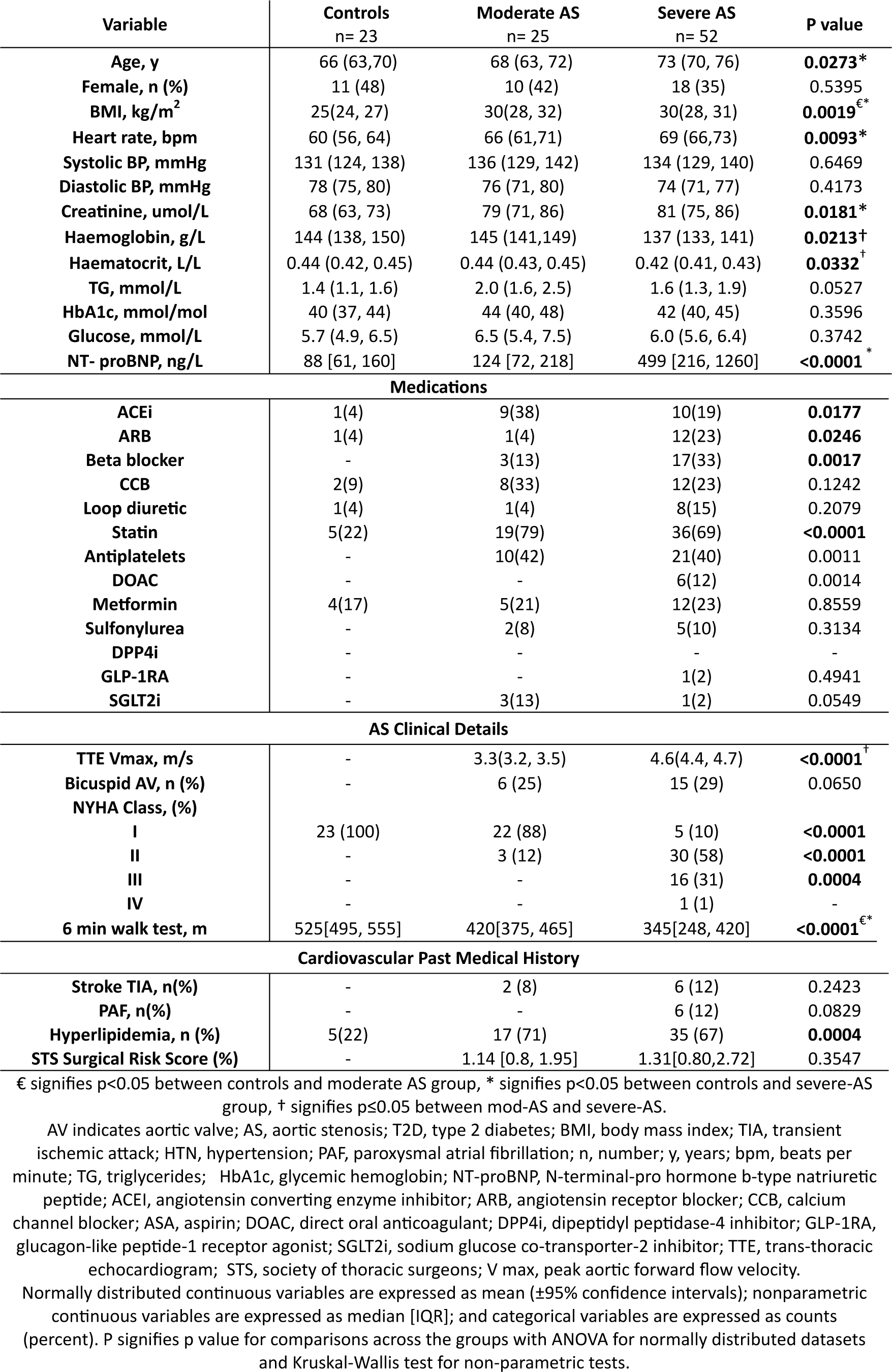
Clinical Characteristics and Biochemistry of All Groups.

| Variable | Controls<br>n= 23 | Moderate AS<br>n= 25 | Severe AS<br>n= 52 | P value |
| --- | --- | --- | --- | --- |
| Age, y | 66 (63,70) | 68 (63, 72) | 73 (70, 76) | **0.0273\*** |
| Female, n (%) | 11 (48) | 10 (42) | 18 (35) | 0.5395 |
| BMI, kg/m<sup>2</sup> | 25(24, 27) | 30(28, 32) | 30(28, 31) | **0.0019<sup>€\*</sup>** |
| Heart rate, bpm | 60 (56, 64) | 66 (61,71) | 69 (66,73) | **0.0093\*** |
| Systolic BP, mmHg | 131 (124, 138) | 136 (129, 142) | 134 (129, 140) | 0.6469 |
| Diastolic BP, mmHg | 78 (75, 80) | 76 (71, 80) | 74 (71, 77) | 0.4173 |
| Creatinine, umol/L | 68 (63, 73) | 79 (71, 86) | 81 (75, 86) | **0.0181\*** |
| Haemoglobin, g/L | 144 (138, 150) | 145 (141,149) | 137 (133, 141) | **0.0213<sup>†</sup>** |
| Haematocrit, L/L | 0.44 (0.42, 0.45) | 0.44 (0.43, 0.45) | 0.42 (0.41, 0.43) | **0.0332<sup>†</sup>** |
| TG, mmol/L | 1.4 (1.1, 1.6) | 2.0 (1.6, 2.5) | 1.6 (1.3, 1.9) | 0.0527 |
| HbA1c, mmol/mol | 40 (37, 44) | 44 (40, 48) | 42 (40, 45) | 0.3596 |
| Glucose, mmol/L | 5.7 (4.9, 6.5) | 6.5 (5.4, 7.5) | 6.0 (5.6, 6.4) | 0.3742 |
| NT- proBNP, ng/L | 88 [61, 160] | 124 [72, 218] | 499 [216, 1260] | **<0.0001<sup>\*</sup>** |
| **Medications** |  |  |  |  |
| ACEi | 1(4) | 9(38) | 10(19) | **0.0177** |
| ARB | 1(4) | 1(4) | 12(23) | **0.0246** |
| Beta blocker | - | 3(13) | 17(33) | **0.0017** |
| CCB | 2(9) | 8(33) | 12(23) | 0.1242 |
| Loop diuretic | 1(4) | 1(4) | 8(15) | 0.2079 |
| Statin | 5(22) | 19(79) | 36(69) | **<0.0001** |
| Antiplatelets | - | 10(42) | 21(40) | 0.0011 |
| DOAC | - | - | 6(12) | 0.0014 |
| Metformin | 4(17) | 5(21) | 12(23) | 0.8559 |
| Sulfonylurea | - | 2(8) | 5(10) | 0.3134 |
| DPP4i | - | - | - | - |
| GLP-1RA | - | - | 1(2) | 0.4941 |
| SGLT2i | - | 3(13) | 1(2) | 0.0549 |
| **AS Clinical Details** |  |  |  |  |
| TTE Vmax, m/s | - | 3.3(3.2, 3.5) | 4.6(4.4, 4.7) | **<0.0001<sup>†</sup>** |
| Bicuspid AV, n (%) | - | 6 (25) | 15 (29) | 0.0650 |
| NYHA Class, (%) |  |  |  |  |
| I | 23 (100) | 22 (88) | 5 (10) | **<0.0001** |
| II | - | 3 (12) | 30 (58) | **<0.0001** |
| III | - | - | 16 (31) | **0.0004** |
| IV | - | - | 1 (1) | - |
| 6 min walk test, m | 525[495, 555] | 420[375, 465] | 345[248, 420] | **<0.0001<sup>€\*</sup>** |
| **Cardiovascular Past Medical History** |  |  |  |  |
| Stroke TIA, n(%) | - | 2 (8) | 6 (12) | 0.2423 |
| PAF, n(%) | - | - | 6 (12) | 0.0829 |
| Hyperlipidemia, n (%) | 5(22) | 17 (71) | 35 (67) | **0.0004** |
| STS Surgical Risk Score (%) | - | 1.14 [0.8, 1.95] | 1.31[0.80,2.72] | 0.3547 |

€ signifies p<0.05 between controls and moderate AS group, \* signifies p<0.05 between controls and severe-AS group, † signifies p≤0.05 between mod-AS and severe-AS.
AV indicates aortic valve; AS, aortic stenosis; T2D, type 2 diabetes; BMI, body mass index; TIA, transient ischemic attack; HTN, hypertension; PAF, paroxysmal atrial fibrillation; n, number; y, years; bpm, beats per minute; TG, triglycerides; HbA1c, glycemic hemoglobin; NT-proBNP, N-terminal-pro hormone b-type natriuretic peptide; ACEi, angiotensin converting enzyme inhibitor; ARB, angiotensin receptor blocker; CCB, calcium channel blocker; ASA, aspirin; DOAC, direct oral anticoagulant; DPP4i, dipeptidyl peptidase-4 inhibitor; GLP-1RA, glucagon-like peptide-1 receptor agonist; SGLT2i, sodium glucose co-transporter-2 inhibitor; TTE, trans-thoracic echocardiogram; STS, society of thoracic surgeons; V max, peak aortic forward flow velocity.
Normally distributed continuous variables are expressed as mean (±95% confidence intervals); nonparametric continuous variables are expressed as median [IQR]; and categorical variables are expressed as counts (percent). P signifies p value for comparisons across the groups with ANOVA for normally distributed datasets and Kruskal-Wallis test for non-parametric tests.

Compared to controls, patients with severe and moderate AS had higher BMI (controls:25[24,27], Mod-AS:30[28,32], Severe-AS:30[28,31]; *P*=0.0019). The 3 groups were matched for blood pressure, fasting glucose and HBA1c; but the resting heart rate was higher, and the haemoglobin was lower in the severe AS group (Table-1).

Patients with severe AS had significantly higher NTproBNP levels than the moderate AS and the control groups (controls:88 [61,160], Mod-AS:124[72,218], Severe-AS:499[216,1260]; *P*<0.0001).

### Cardiovascular magnetic resonance and ^31^phosphorus-magnetic resonance spectroscopy findings

#### Cardiac remodeling

Compared to controls, both AS groups showed LV concentric remodeling with a stepwise increase from the moderate to severe AS groups in the LV mass-index (controls:46[40,51], Mod-AS:58[51,65], Severe-AS:70[65,75]g/m^2^; *P*<0.0001) (Figure-2A), LV concentricity index (LV mass to LV end-diastolic volume) (controls:0.58[0.54,0.62], Mod-AS:0.74[0.64,0.84], Severe-AS:0.89[0.83,0.95]g/mL; *P*<0.0001] (Figure-2B), and LV wall thickness (controls:9.9[9.2,10.5], Mod-AS:12.3[11.2,13.3], Severe-AS:14.8[14.1,15.5]; *P*<0.0001) with significant differences also between the moderate and severe AS groups. However, the LV mass (controls:87[74,100], Mod-AS:103[86,120], Severe-AS:133[121,145]g; *P*<0.0001) was only significantly increased in patients with severe AS compared to both the Mod-AS and the control groups.

**Figure 2:**
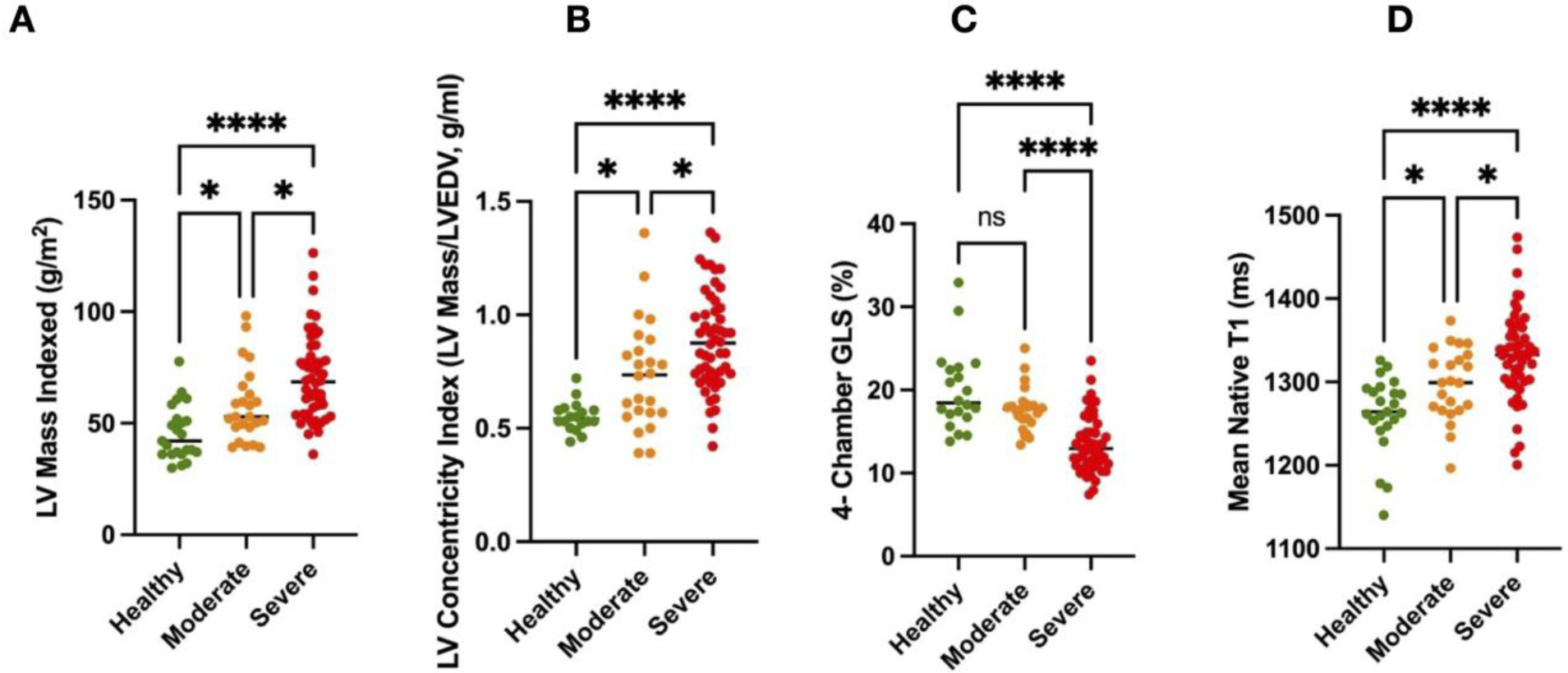
Column scatter graphs with means and 95% confidence intervals showing differences in (A) Left ventricular (LV) mass indexed to the body surface area (g/m^2^), (B) LV concentricity index (LV mass over LV end-diastolic volume ratio, g/ml), (C) Global longitudinal shortening (%), (D) Mean native T1 (miliseconds) between the patients with moderate AS, severe AS and controls.

In line with the recruitment criteria, LV ejection fraction was normal in all patients with moderate or severe AS with no significant differences across the study groups (Table-2). However, patients with severe AS showed significantly impaired GLS (controls:19.9[17.6,22.2], Mod-AS:17.7[16.6,18.8], Severe-AS:13.4[12.5,14.4]%; *P*<0.0001) (Figure-2C) and left atrial (LA) function (controls:57[52,63], Mod-AS:59[52,66], Severe-AS:40[35,44]%; *P*<0.0001) compared to both the moderate AS group and the controls.

**Table 2:**
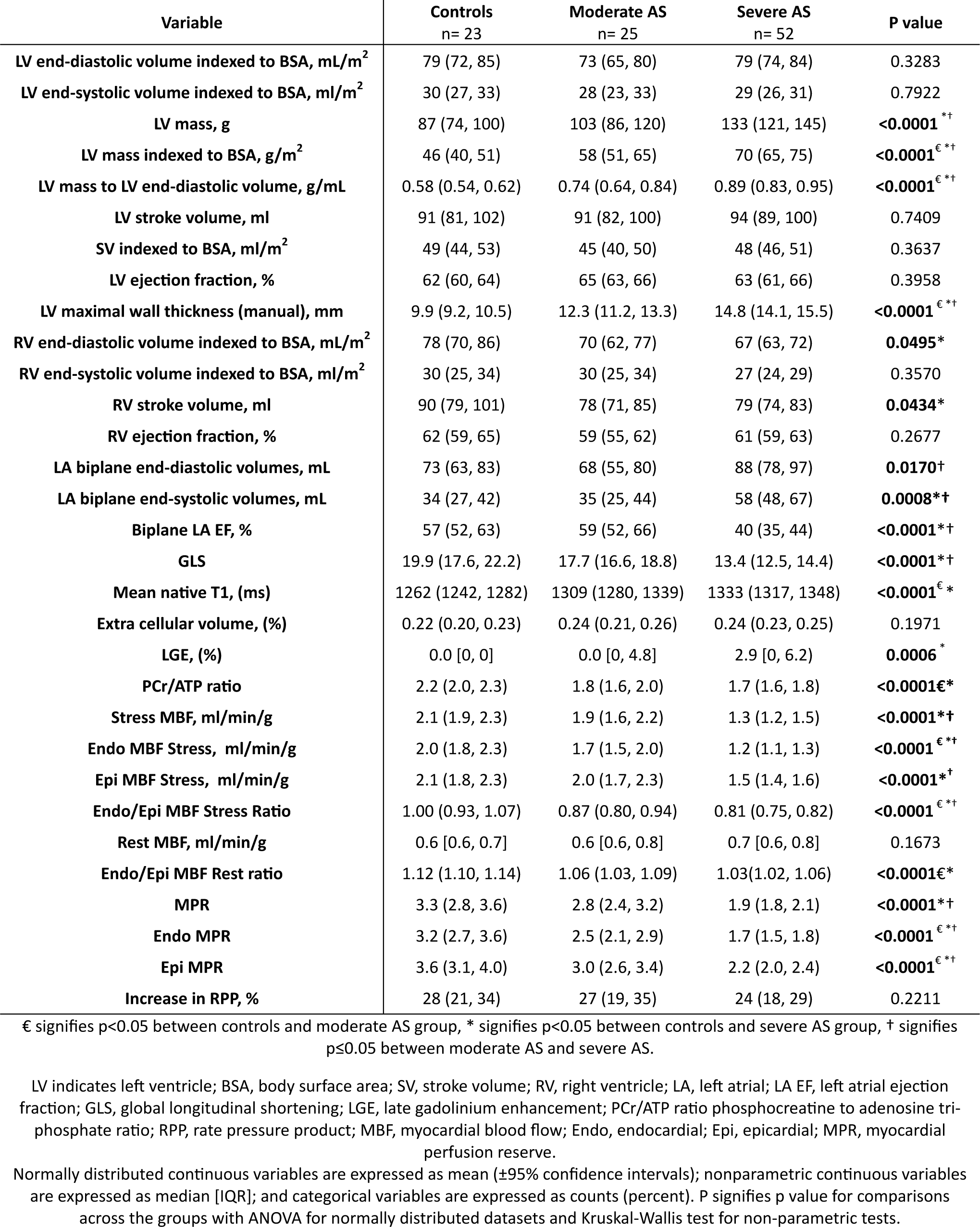
CMR and ^31^P-MRS findings of All Groups.

Only the severe AS group showed significant myocardial scarring on LGE (2.9[0,6.2]%). The native T1 was higher in both AS groups compared to controls, with significant differences also between the moderate and severe AS groups (controls:1262[1242,1282], Mod-AS:1309[1280,1339], Severe-AS:1333[1317,1348]; *P*<0.0001) (Figure-2D). There was no significant difference in ECV fraction across the groups (controls: 0.22[0.20,0.23], Mod-AS:0.24[0.21,0.26], Severe-AS:0.24[0.23,0.25]%; *P*=0.20), suggesting the increase in T1 was reflective of cellular hypertrophy as opposed to interstitial fibrosis in the AS cohorts,.

The AS severity (peak aortic valve velocity [Vmax]) correlated with the LV-mass (r=0.42, *P*=0.0005, Figure-5A) and GLS (r=-0.47, *P*=0.0001, Figure-5B). There were similar correlations detected with aortic valve area by planimetry with the LV mass and LV mass-index and GLS (supplementary materials figure 2).

Figure 6 shows representative adenosine-stress myocardial perfusion map examples from a patient with moderate aortic stenosis and a patient with severe aortic stenosis.

#### Cardiac energetics and perfusion

Compared to controls, both AS groups showed significantly lower myocardial PCr/ATP ratio (controls:2.2[2.0,2.5], Mod-AS:1.8[1.6,2.0], Severe-AS:1.7[1.6,1.8]; *P*<0.0001) with no significant difference between the moderate and severe AS groups (Figure-3). There was a negative correlation between the PCr/ATP and the LV mass (r=-0.30, *P=*0.008), but not LV mass-index.

**Figure 3:**
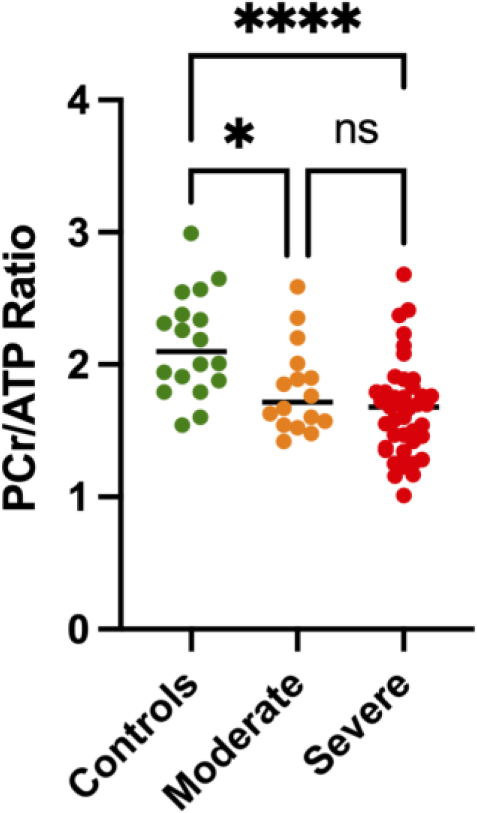
Column scatter graphs with means and 95% confidence intervals showing differences in myocardial phosphocreatine to adenosine triphosphate ratio (PCr/ATP) between the patients with moderate aortic stenosis (AS), severe AS and controls.

Only the severe AS group showed significant reductions in global adenosine-stress MBF (controls:2.1[1.9,2.3], Mod-AS:1.9[1.6,2.2], Severe-AS:1.3[1.2,1.5]ml/min/g; *P*<0.0001) (Figure-4A) and global MPR (controls:3.3[2.8,3.6], Mod-AS:2.8[2.4,3.2], Severe-AS:1.9[1.8,2.1]; *P*<0.0001) (Figure-4E).

**Figure 4:**
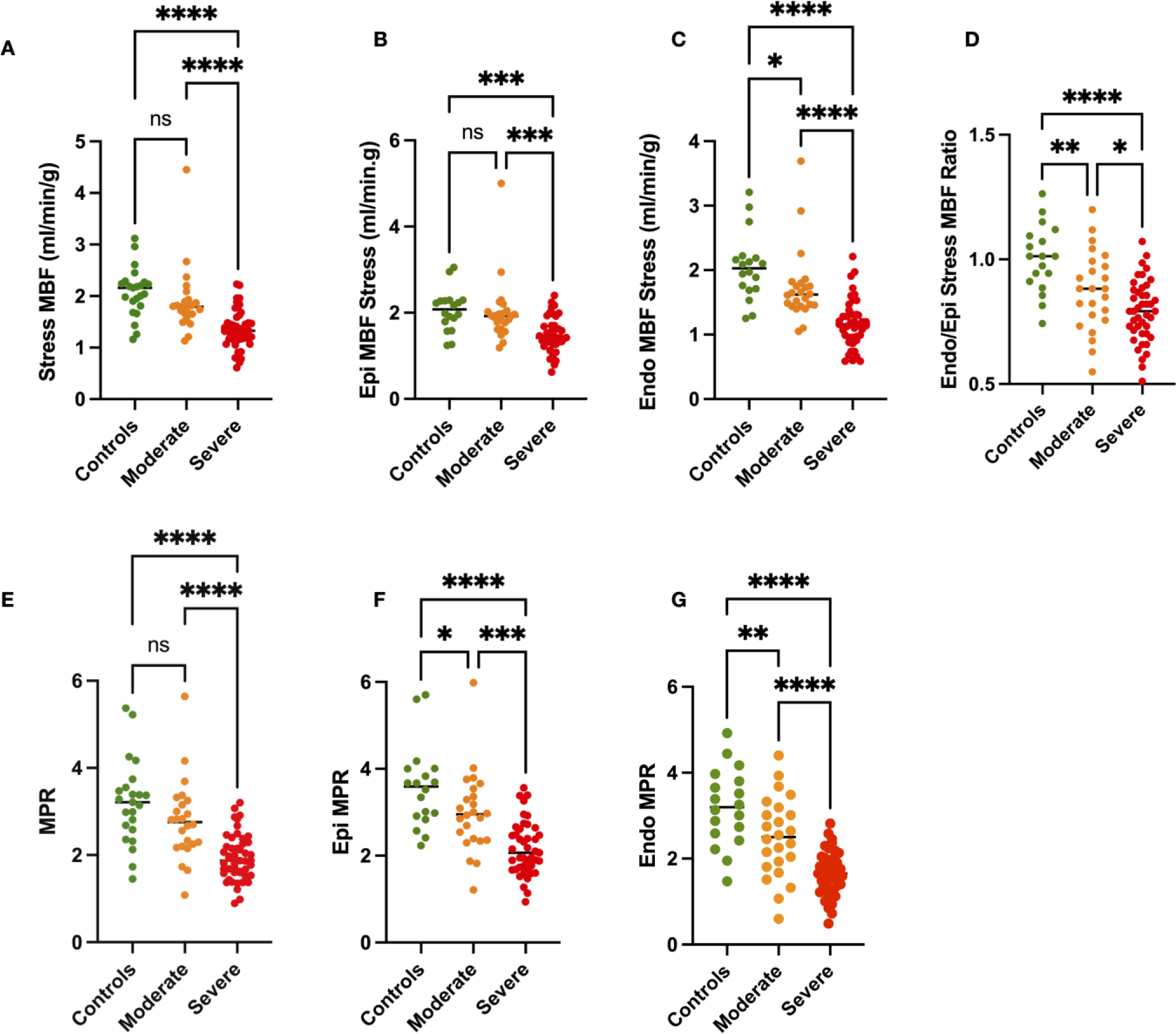
Column scatter graphs with means and 95% confidence intervals showing differences in (A) phosphocreatine to ATP ratio (PCr/ATP), (B) Vasodilator stress myocardial blood flow (MBF, ml/min/g), (C) myocardial perfusion reserve (MPR) between the patients with moderate AS, severe AS and controls.

However, adenosine-stress Endo-MBF (controls: 2.0 [1.8, 2.3], Mod-AS: 1.7 [1.5,2.0], Severe-AS: 1.2 [1.1, 1.3] ml/min/g; *P*<0.0001) (Figure-4C), adenosine-stress Endo/Epi ratio (controls:1.00[0.93,1.07], Mod-AS:0.87[0.80,0.94], Severe-AS:0.81[0.75,0.82]; *P<*0.0001), (Figure-4D), Epi-MPR (controls: 3.6 [3.1, 4.0], Mod-AS: 3.0 [2.6, 3.4], Severe-AS: 2.2 [2.0, 2.4]; *P<*0.0001) (Figure-4F), Endo-MPR (controls:3.2 [2.7, 3.6], Mod-AS: 2.5 [2.1, 2.9], Severe-AS: 1.7 [1.5, 1.8]; *P<*0.0001) (Figure-4G) and rest-Endo/Epi (controls:1.12[1.10,1.14], Mod-AS:1.06[1.03,1.09], Severe-AS:1.03[1.02,1.06]; *P*<0.0001) were all significantly reduced in both Mod-AS and Severe-AS. Among the layer-specific perfusion metrics only Epi-MBF was not significantly reduced in Mod-AS but only in Severe-AS (controls: 2.1 [1.8, 2.3], Mod-AS: 2.0 [1.7, 2.3], Severe-AS: 1.5 [1.4, 1.6] ml/min/g; *P*<0.0001).

There were significant correlations between the AS severity (Vmax on TTE) and the global-global MPR (r=-0.44; *P*=0.0005) (Figure-5C), Endo-MPR (r=0.1852; *P*=0.0007) (Figure-5D), stress-MBF (r=-0.45; *P*=0.0003) (Figure-5E) and adenosine-stress Endo-MBF (r=0.2746; P<0.0001) (Figure-5F). In parallel, there were also significant correlations detected with aortic valve area (cm^2^) by planimetry with the perfusion metrics (supplementary materials figure 2).

**Figure 5:**
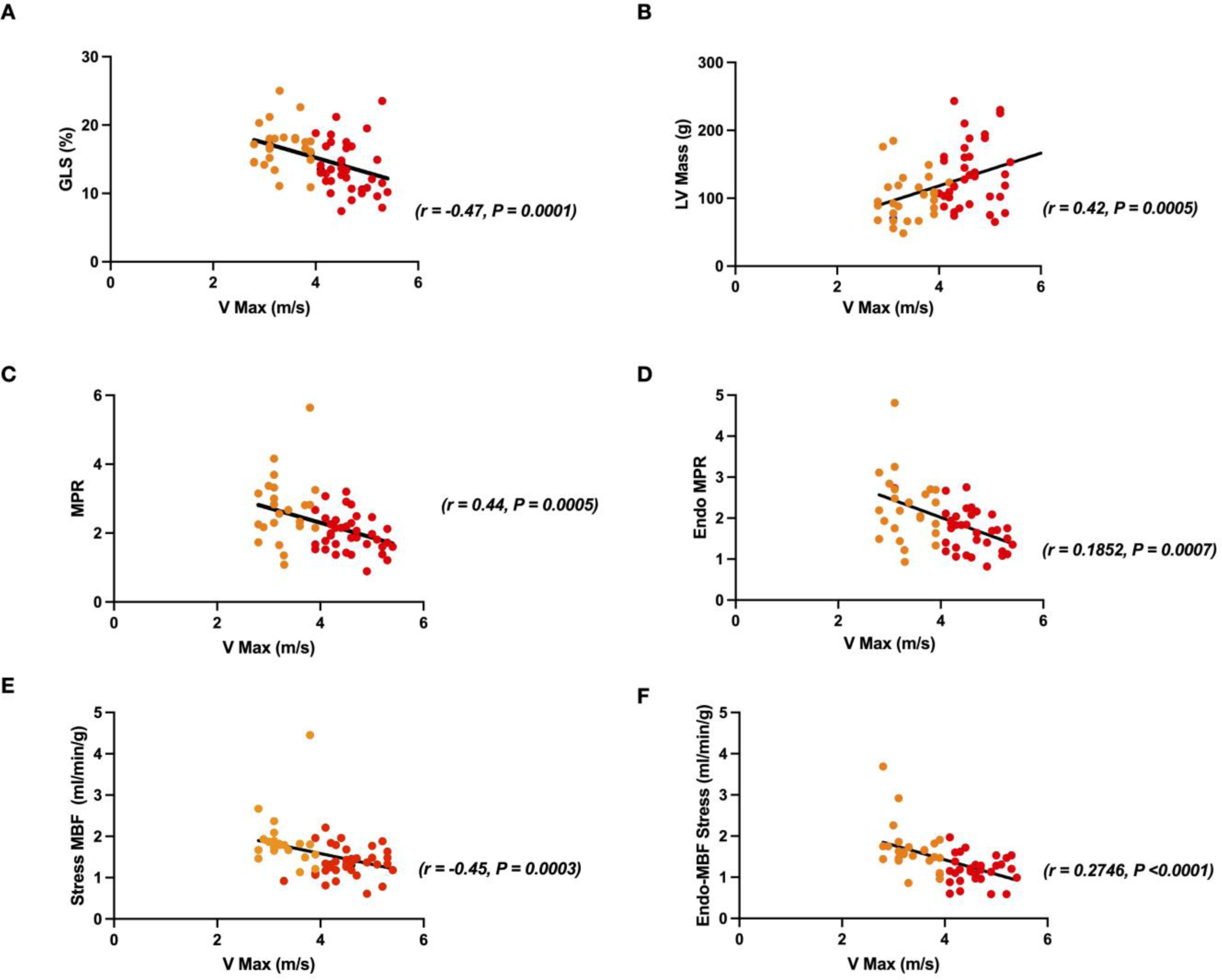
Scatterplots showing the correlations of the peak aortic valve velocity (Vmax, m/s) severity of aortic stenosis with: (A) global longitudinal shortening (GLS, %), (B) Left ventricular mass (LV Mass, g), (C) Myocardial perfusion reserve (MPR), D) Endocardial (endo) Myocardial perfusion reserve (MPR), (E) Vasodilator stress myocardial blood flow (MBF, ml/min/g) and (F) Vasodilator endocardial (Endo) stress myocardial blood flow (MBF, ml/min/g). Orange dots represent patients with moderate AS and red dots represent patients with severe AS. P signifies p value for comparisons between each of the variables when measured against peak aortic valve velocity. The r value represents correlation coefficient as measured by linear regression analysis.

**Figure 6:**
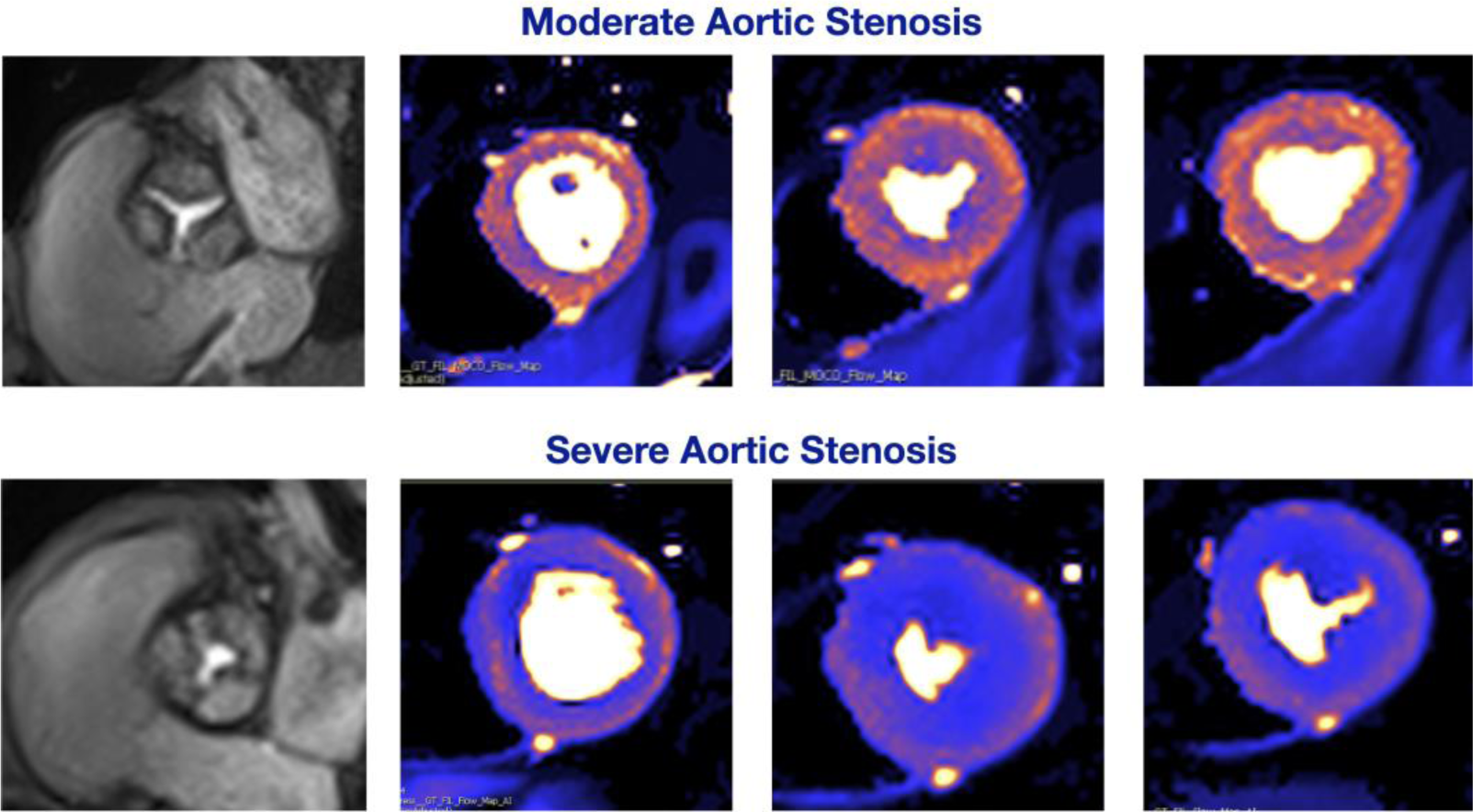
Representative adenosine-stress myocardial perfusion map examples from a patient with moderate aortic stenosis (top row) and a patient with severe aortic stenosis (bottom row).

## DISCUSSION

Given the relevance of myocardial health in the prognosis of patients with AS, this study prospectively compared myocardial remodelling, energetics and perfusion parameters in patients with moderate and severe AS with preserved LV ejection fraction against controls without AS^7,11,25^. Moderate and severe AS were both associated with cardiac concentric hypertrophy, myocardial energetic impairment, and reduced exercise distance on 6-minute walk test. While moderate AS was not associated with reductions in the global myocardial perfusion metrics of rest and stress MBF or MPR, the transmural perfusion dynamics were significantly affected, with significant reductions in stress Endo-MBF, Endo/Epi-MBF gradient, and Endo-and Epi-MPR compared to controls. Severe AS was associated with global as well as endocardial and epicardial reductions in MBF and MPR, and reductions in the Endo/Epi-MBF gradient values. However, there was a stepwise worsening of the hypertrophic remodeling of the LV with progression of the severity of the valve stenosis. The patients with moderate AS did not exhibit significant reductions in LV contractile function, nor did they show evidence of myocardial scarring, when these findings were evident in patients with severe AS.

### Cardiac remodeling

In line with the findings from this study, a previous longitudinal surveillance study of patients with AS showed the main phenotypic change with progression of AS severity to be the development of concentric LV hypertrophy.^26^ While milder compared to that seen in severe AS, the degree of adverse myocardial remodeling associated with moderate AS in this study may be clinically important for the cardiovascular clinical outcomes in patients with moderate AS. There is consistent evidence strongly linking LV concentric remodeling and hypertrophy to adverse cardiovascular outcomes^27–29^. An adverse association between greater LV mass-index and worse clinical outcomes was shown among patients with severe and moderate AS^30^.

The remodeling of the LV in AS involves the process of cardiomyocyte hypertrophy and extracellular matrix expansion to normalize the wall tension and maintain cardiac output and eventually interstitial fibrosis and scarring^31^. The lack of myocardial scarring in moderate AS group in this study in particular may be of significant clinical relevance because among the imaging biomarkers of ventricular decompensation in AS, myocardial scarring on CMR has consistently been shown to be the strongest associate of all-cause and cardiovascular mortality after AVR^7,25,32^.

In this study, the native T1 values were significantly elevated in both AS groups, increasing with the rising stenosis severity. However, the T1 elevation was in isolation, without an accompanying significant increase in the ECV fraction even in the severe AS group. This finding is also consistent with a previous study showing that ECV values do not change with the severity of AS^33^. The normal-range ECV values even in the severe AS category may potentially be reflective of the low surgical risk score (Society of Thoracic Surgeons risk score<1%) of the severe AS group and may suggest cardiomyocyte hypertrophy without myocardial fibrosis^11,34^. In AS, myocardial fibrosis is a key pathological process driving the transition from hypertrophy to heart failure. In a study using invasive myocardial biopsy and CMR imaging, myocardial fibrosis in severe AS was shown to have three main alterations: endocardial thickening, subendocardial microscars, and diffuse interstitial fibrosis^35^. This landmark study demonstrated that neither histological collagen volume fraction nor the CMR parameters ECV and LGE capture this fibrosis in its totality. This may provide a potential explanation for the discordance between the LGE findings showing scar in the severe AS group while the ECV remains within the normal range.

### Cardiac energetics and perfusion

At metabolic level, LV hypertrophy is associated with decreased myocardial high-energy phosphate pool (energetics) and reductions in the creatine kinase flux^36^. These changes in turn cause myocardial energetic starvation in patients with severe AS^16^. Eventually, this energy-starved, sub-optimally perfused myocardial state causes myocardial ischemia, cardiomyocyte death, irreversible myocardial scarring, and even sudden death^15,37^. Although the momentum of these changes varies between patients, the progression of the valve restriction and aggravation of cardiac hypertrophy eventually results in development of symptoms, HF and death^31^. However, the level of energetic deficit was similar between the moderate and severe AS groups and this correlated with the LV mass. This suggests that even a milder degree of hypertrophy is associated with a decreased myocardial high-energy phosphate pool.

To the best of our knowledge, this is the first report of global and transmural myocardial perfusion parameters using automated, in-line pixelwise quantitative myocardial perfusion CMR in patients with moderate AS. Myocardial hypertrophy is accompanied by capillary density reduction, microcirculatory dysfunction and impaired stress MBF^38,39^. The results from this study confirm the previous studies showing significant reductions in global stress MBF in severe AS, whilst also highlighting that the extent of the impaired myocardial perfusion is subject to the severity of AS. In normal hearts, resting MBF is the greatest in the endocardium (Endo/Epi ratio >1), but epicardial MBF is augmented during adenosine-induced hyperemia to a greater extent. Prior investigations have consistently reported that in AS, preferential coronary flow shifts from the endocardium to epicardium results in a significant decrease in endocardial MBF^13^. The reversal of normal Endo/Epi blood flow ratio is fundamental to the pathophysiology of AS, resulting in subendocardial ischemia, apoptosis, and fibrosis—with clinical manifestation as angina despite non-obstructed epicardial coronary arteries. This study shows for the first time that myocardial layer-specific perfusion parameters are already significantly affected in moderate AS.

### Clinical perspectives

Underscoring the importance of myocardial health in the prognosis, patients with AS even after timely AVR remain at higher risk of cardiovascular mortality (Hazard Ratio [HR] 1.79 [95% CI: 1.75-1.83])^40,41^, which is primarily associated with heart failure^42,43^. Studies of early intervention strategies are currently proceeding to address the issue of excess cardiovascular death in patients with AS compared with the reference population even after undergoing AVR^40,41^. These trials propose that early intervention can prevent irreversible myocardial pre-AVR damage and will therefore prevent adverse post-AVR outcomes, as myocardial health governs post-AVR prognosis. Therefore, the paradigm for treatment of AS might be shifting from the focus on valvular obstruction to extra-valvular myocardial damage.

The accumulating evidence from small-sized randomized clinical trials^44,45^ supports further expansion of indications for earlier AVR for patients with AS before the onset of symptoms to prevent myocardial decompensation, and several large trials are currently underway testing this hypothesis^46^. In particular, there is ongoing work to investigate whether myocardial scar can be used to guide the intervention timings of AS patients.^47^ A separate large trial is comparing the efficacy of early transcatheter AVR strategy to that of clinical surveillance for patients with moderate AS on reducing major adverse cardiac events^48^.

However, the evidence of worse outcomes even in moderate AS may suggest there are other important prognostic factors at play that are evident even in less than severe AS. This study contributes to the existing literature by showing that while milder compared to that seen in severe AS, the degree of adverse myocardial remodeling associated with moderate AS may be clinically important for the cardiovascular clinical outcomes in patients with moderate AS. Larger prospective serial studies and randomized trials are needed to better understand the mechanisms of the high cardiovascular and all-cause mortality rates in patients with moderate AS.

## LIMITATIONS

This study had several limitations. Due to the cross-sectional nature of the study causality of the observed differences cannot be inferred and the small sample recruited at a single site increases the risk of bias and type I error. While obstructive coronary artery stenosis was excluded using X-ray coronary angiography in all patients with severe AS, the control group and the moderate AS group did not undergo anatomical coronary imaging. Significant coronary artery disease was deemed to be unlikely in these cohorts supported by the absence of both myocardial infarction and regional stress perfusion defects on their imaging studies.

A larger study with a longer follow-up duration will be required to confirm the clinical significance of the cardiac remodeling findings to the high cardiovascular and all-cause mortality rates in patients with moderate AS. Finally, the complexity of the imaging protocol, in particular the MR spectroscopy, may limit its widespread use.

## CONCLUSIONS

Moderate and severe AS are both associated with cardiac concentric hypertrophy, reductions in myocardial energetics, subendocardial hypoperfusion, and limitations in exercise distance. Patients with Severe-AS exhibit a more pronounced phenotype with worse LV hypertrophy, contractile dysfunction and myocardial scarring compared to patients with Mod-AS.

## Funding

The study was jointly supported by the Wellcome Trust (grant number: 221690/Z/20/Z) and Diabetes UK (grant number:18/0005870).

EL acknowledges support from the Wellcome Trust Clinical Career Development Fellowship (grant number: 221690/Z/20/Z), Diabetes UK (grant number:18/0005870) and National Institute for Health and Care Research (NIHR) Leeds Biomedical Research Centre. LV is funded by a Sir Henry Dale Fellowship supported jointly by the Wellcome Trust and the Royal Society (#221805/Z/20/Z), and he also acknowledges the support of the Slovak Grant Agencies VEGA (#2/0003/20) and APVV (#19-0032). Funding for open access charge: Wellcome Trust (grant number: 221690/Z/20/Z).

### ABBREVIATIONS

|  |  |  |
| --- | --- | --- |
| 111 | ACEi | Angiotensin converting enzyme inhibitor |
| 112 | ADP | Adenosine diphosphate |
| 113 | AF | Atrial fibrillation |
| 114 | AS | Aortic stenosis |
| 115 | ATP | Adenosine triphosphate |
| 116 | AVR | Aortic valve replacement |
| 117 | AW | Acquisition Weighting |
| 118 | BMI | Body mass index |
| 119 | CK | Creatine kinase |
| 120 | CMR | Cardiovascular magnetic resonance |
| 121 | CSI | Chemical shift imaging |
| 122 | CV | Cardiovascular |
| 123 | ECV | Extra cellular volume fraction |
| 124 | ESC | European Society of Cardiology |
| 125 | FFA | Free fatty acids |
| 126 | LGE | Late gadolinium enhancement |
| 127 | LV | Left ventricle |
| 128 | Mod-AS | Moderate aortic stenosis |
| 129 | MOLLI | Modified look locker inversion recovery |
| 130 | NT-proBNP | N-terminal pro hormone B-type natriuretic peptide |
| 131 | NYHA | New York Heart Association |
| 132 | <sup>31</sup>P-MRS | <sup>31</sup> Phosphorus magnetic resonance spectroscopy |
| 133 | PCr | Phosphocreatine |
| 134 | RV | Right ventricle |
| 135 | Severe-AS | Severe aortic stenosis |
| 136 | SSFP | Steady State Free Precession |
| 137 | SAVR | Surgical aortic valve replacement |
| 138 | SV | Stroke volume |
| 139 | TAVR | Transcatheter aortic valve replacement |

## Data Availability

The data will be shared on reasonable request to the corresponding author.

## Acknowledgements

Author-specific contributions to the study are as follows: MG and HP contributed to patient recruitment, data acquisition, analysis, and interpretation, drafting of the manuscript and revisions. AC, ST, NJ and SK contributed to data analysis, interpretation, and manuscript revision. RC, LDR, PS, SP, MRD, and JPG contributed to data interpretation and manuscript revision. HX, LV, PK provided essential support for the scan sequences and manuscript revision. EL contributed to study conception and design, funding, data acquisition, analysis, and interpretation, drafting of the manuscript, revisions, and study supervision. All authors approved the final version of the manuscript. For the purpose of Open Access, the authors will apply a CC BY public copyright license to any Author Accepted Manuscript version arising from this submission. The funding bodies played no role in the design of the study and collection, analysis, and interpretation of data and in writing the manuscript. The views expressed are those of the author(s) and not necessarily those of the NHS, the NIHR or the Department of Health and Social Care.

## References

1. Nishimura RA, Otto CM, Bonow RO, Carabello BA, Erwin JP, 3rd, Fleisher LA, Jneid H, Mack MJ, McLeod CJ, O’Gara PT, et al. 2017 AHA/ACC Focused Update of the 2014 AHA/ACC Guideline for the Management of Patients With Valvular Heart Disease: A Report of the American College of Cardiology/American Heart Association Task Force on Clinical Practice Guidelines. Circulation. 2017;135:e1159-e1195. doi: 10.1161/cir.0000000000000503

2. Badiani S, Bhattacharyya S, Aziminia N, Treibel TA, Lloyd G. Moderate Aortic Stenosis: What is it and When Should We Intervene? Interv Cardiol. 2021;16:e09. doi: 10.15420/icr.2021.04

3. Pankayatselvan V, Raber I, Playford D, Stewart S, Strange G, Strom JB. Moderate aortic stenosis: culprit or bystander? Open Heart. 2022;9. doi: 10.1136/openhrt-2021-001743

4. Strange G, Stewart S, Celermajer D, Prior D, Scalia GM, Marwick T, Ilton M, Joseph M, Codde J, Playford D. Poor Long-Term Survival in Patients With Moderate Aortic Stenosis. Journal of the American College of Cardiology. 2019;74:1851–1863. doi: 10.1016/j.jacc.2019.08.004

5. Delesalle G, Bohbot Y, Rusinaru D, Delpierre Q, Maréchaux S, Tribouilloy C. Characteristics and Prognosis of Patients With Moderate Aortic Stenosis and Preserved Left Ventricular Ejection Fraction. Journal of the American Heart Association. 2019;8:e011036. doi: 10.1161/JAHA.118.011036

6. Stewart S, Chan Y-K, Playford D, Strange GA. Incident aortic stenosis in 49 449 men and 42 229 women investigated with routine echocardiography. Heart. 2022;108:875. doi: 10.1136/heartjnl-2021-319697

7. Musa TA, Treibel TA, Vassiliou VS, Captur G, Singh A, Chin C, Dobson LE, Pica S, Loudon M, Malley T, et al. Myocardial Scar and Mortality in Severe Aortic Stenosis. Circulation. 2018;138:1935–1947. doi: 10.1161/CIRCULATIONAHA.117.032839

8. Thornton GD, Musa TA, Rigolli M, Loudon M, Chin C, Pica S, Malley T, Foley JRJ, Vassiliou VS, Davies RH, et al. Association of Myocardial Fibrosis and Stroke Volume by Cardiovascular Magnetic Resonance in Patients With Severe Aortic Stenosis With Outcome After Valve Replacement: The British Society of Cardiovascular Magnetic Resonance AS700 Study. JAMA Cardiol. 2022;7:513–520. doi: 10.1001/jamacardio.2022.0340

9. Rosenhek R, Klaar U, Schemper M, Scholten C, Heger M, Gabriel H, Binder T, Maurer G, Baumgartner H. Mild and moderate aortic stenosis. Natural history and risk stratification by echocardiography. Eur Heart J. 2004;25:199–205. doi: 10.1016/j.ehj.2003.12.002

10. Iung B, Delgado V, Rosenhek R, Price S, Prendergast B, Wendler O, De Bonis M, Tribouilloy C, Evangelista A, Bogachev-Prokophiev A, et al. Contemporary Presentation and Management of Valvular Heart Disease: The EURObservational Research Programme Valvular Heart Disease II Survey. Circulation. 2019;140:1156–1169. doi: 10.1161/circulationaha.119.041080

11. Everett RJ, Treibel TA, Fukui M, Lee H, Rigolli M, Singh A, Bijsterveld P, Tastet L, Musa TA, Dobson L, et al. Extracellular Myocardial Volume in Patients With Aortic Stenosis. J Am Coll Cardiol. 2020;75:304–316. doi: 10.1016/j.jacc.2019.11.032

12. Kellman P, Hansen MS, Nielles-Vallespin S, Nickander J, Themudo R, Ugander M, Xue H. Myocardial perfusion cardiovascular magnetic resonance: optimized dual sequence and reconstruction for quantification. Journal of Cardiovascular Magnetic Resonance. 2017;19:43. doi: 10.1186/s12968-017-0355-5

13. McConkey HZR, Marber M, Chiribiri A, Pibarot P, Redwood SR, Prendergast BD. Coronary Microcirculation in Aortic Stenosis. Circulation: Cardiovascular Interventions.12:e007547. doi: 10.1161/CIRCINTERVENTIONS.118.007547

14. Markley R, Del Buono MG, Mihalick V, Pandelidis A, Trankle C, Jordan JH, Decamp K, Winston C, Carbone S, Billingsley H, et al. Abnormal left ventricular subendocardial perfusion and diastolic function in women with obesity and heart failure and preserved ejection fraction. Int J Cardiovasc Imaging. 2023;39:811–819. doi: 10.1007/s10554-022-02782-x

15. Neubauer S. The Failing Heart — An Engine Out of Fuel. New England Journal of Medicine. 2007;356:1140–1151. doi: 10.1056/NEJMra063052

16. Peterzan Mark A, Clarke William T, Lygate Craig A, Lake Hannah A, Lau Justin YC, Miller Jack J, Johnson E, Rayner Jennifer J, Hundertmark Moritz J, Sayeed R, et al. Cardiac Energetics in Patients With Aortic Stenosis and Preserved Versus Reduced Ejection Fraction. Circulation. 2020;141:1971–1985. doi: 10.1161/CIRCULATIONAHA.119.043450

17. Delesalle G, Bohbot Y, Rusinaru D, Delpierre Q, Marechaux S, Tribouilloy C. Characteristics and Prognosis of Patients With Moderate Aortic Stenosis and Preserved Left Ventricular Ejection Fraction. Journal of the American Heart Association. 2019;8(6) (no pagination).

18. Howard T, Majmundar M, Sarin S, Kumar A, Ajay A, Krishnaswamy A, Reed GW, Harb SC, Harmon E, Dykun I, et al. Predictors of Major Adverse Cardiovascular Events in Patients with Moderate Aortic Stenosis: Implications for Aortic Valve Replacement. Circulation: Cardiovascular Imaging. 2023;16(7):557–565.

19. Casanova C, Celli BR, Barria P, Casas A, Cote C, Torres JPd, Jardim J, Lopez MV, Marin JM, Oca MMd, et al. The 6-min walk distance in healthy subjects: reference standards from seven countries. European Respiratory Journal. 2011;37:150. doi: 10.1183/09031936.00194909

20. Thirunavukarasu S, Jex N, Chowdhary A, Hassan IU, Straw S, Craven TP, Gorecka M, Broadbent D, Swoboda P, Witte KK, et al. Empagliflozin Treatment Is Associated With Improvements in Cardiac Energetics and Function and Reductions in Myocardial Cellular Volume in Patients With Type 2 Diabetes. Diabetes. 2021;70:2810–2822. doi: 10.2337/db21-0270

21. Chowdhary A, Thirunavukarasu S, Jex N, Coles L, Bowers C, Sengupta A, Swoboda P, Witte K, Cubbon R, Xue H, et al. Coronary microvascular function and visceral adiposity in patients with normal body weight and type 2 diabetes. Obesity. 2022;30:1079–1090. doi: 10.1002/oby.23413

22. Cerqueira MD, Weissman NJ, Dilsizian V, Jacobs AK, Kaul S, Laskey WK, Pennell DJ, Rumberger JA, Ryan T, Verani MS, et al. Standardized myocardial segmentation and nomenclature for tomographic imaging of the heart. A statement for healthcare professionals from the Cardiac Imaging Committee of the Council on Clinical Cardiology of the American Heart Association. Circulation. 2002;105:539–542. doi: 10.1161/hc0402.102975

23. Kellman P, Hansen MS, Nielles-Vallespin S, Nickander J, Themudo R, Ugander M, Xue H. Myocardial perfusion cardiovascular magnetic resonance: optimized dual sequence and reconstruction for quantification. J Cardiovasc Magn Reson. 2017;19:43. doi: 10.1186/s12968-017-0355-5

24. Jex N, Chowdhary A, Thirunavukarasu S, Procter H, Sengupta A, Natarajan P, Kotha S, Poenar AM, Swoboda P, Xue H, et al. Coexistent Diabetes Is Associated With the Presence of Adverse Phenotypic Features in Patients With Hypertrophic Cardiomyopathy. Diabetes Care. 2022;45:1852–1862. doi: 10.2337/dc22-0083

25. Thornton GD, Vassiliou VS, Musa TA, Aziminia N, Craig N, Dattani A, Davies RH, Captur G, Moon JC, Dweck MR, et al. Myocardial Scar and Remodelling Predict Long-Term Mortality in Severe Aortic Stenosis Beyond 10 Years. European Heart Journal. 2024:ehae067. doi: 10.1093/eurheartj/ehae067

26. Stassen J, Ewe SH, Pio SM, Pibarot P, Redfors B, Leipsic J, Genereux P, Van Mieghem NM, Kuneman JH, Makkar R, et al. Managing Patients With Moderate Aortic Stenosis. JACC Cardiovasc Imaging. 2023;16:837–855. doi: 10.1016/j.jcmg.2022.12.013

27. Bluemke DA, Kronmal RA, Lima JAC, Liu K, Olson J, Burke GL, Folsom AR. The Relationship of Left Ventricular Mass and Geometry to Incident Cardiovascular Events: The MESA Study. Journal of the American College of Cardiology. 2008;52:2148–2155. doi: 10.1016/j.jacc.2008.09.014

28. Bang CN, Gerdts E, Aurigemma GP, Boman K, de Simone G, Dahlöf B, Køber L, Wachtell K, Devereux RB. Four-Group Classification of Left Ventricular Hypertrophy Based on Ventricular Concentricity and Dilatation Identifies a Low-Risk Subset of Eccentric Hypertrophy in Hypertensive Patients. Circulation: Cardiovascular Imaging. 2014;7:422–429. doi: 10.1161/CIRCIMAGING.113.001275

29. Koren MJ, Devereux RB, Casale PN, Savage DD, Laragh JH. Relation of Left Ventricular Mass and Geometry to Morbidity and Mortality in Uncomplicated Essential Hypertension. Annals of Internal Medicine. 1991;114:345–352. doi: 10.7326/0003-4819-114-5-345

30. Gonzales H, Douglas PS, Pibarot P, Hahn RT, Khalique OK, Jaber WA, Cremer P, Weissman NJ, Asch FM, Zhang Y, et al. Left Ventricular Hypertrophy and Clinical Outcomes Over 5 Years After TAVR: An Analysis of the PARTNER Trials and Registries. JACC: Cardiovascular Interventions. 2020;13:1329–1339. doi: 10.1016/j.jcin.2020.03.011

31. Everett RJ, Clavel M-A, Pibarot P, Dweck MR. Timing of intervention in aortic stenosis: a review of current and future strategies. Heart. 2018;104:2067. doi: 10.1136/heartjnl-2017-312304

32. Dweck MR, Joshi S, Murigu T, Alpendurada F, Jabbour A, Melina G, Banya W, Gulati A, Roussin I, Raza S, et al. Midwall Fibrosis Is an Independent Predictor of Mortality in Patients With Aortic Stenosis. Journal of the American College of Cardiology. 2011;58:1271–1279. doi: 10.1016/j.jacc.2011.03.064

33. Chin CWL, Everett RJ, Kwiecinski J, Vesey AT, Yeung E, Esson G, Jenkins W, Koo M, Mirsadraee S, White AC, et al. Myocardial Fibrosis and Cardiac Decompensation in Aortic Stenosis. JACC Cardiovasc Imaging. 2017;10:1320–1333. doi: 10.1016/j.jcmg.2016.10.007

34. Swoboda PP, McDiarmid AK, Erhayiem B, Ripley DP, Dobson LE, Garg P, Musa TA, Witte KK, Kearney MT, Barth JH, et al. Diabetes Mellitus, Microalbuminuria, and Subclinical Cardiac Disease: Identification and Monitoring of Individuals at Risk of Heart Failure. Journal of the American Heart Association: Cardiovascular and Cerebrovascular Disease. 2017;6:e005539. doi: 10.1161/JAHA.117.005539

35. Treibel TA, López B, González A, Menacho K, Schofield RS, Ravassa S, Fontana M, White SK, DiSalvo C, Roberts N, et al. Reappraising myocardial fibrosis in severe aortic stenosis: an invasive and non-invasive study in 133 patients. Eur Heart J. 2018;39:699–709. doi: 10.1093/eurheartj/ehx353

36. Bottomley PA, Wu KC, Gerstenblith G, Schulman SP, Steinberg A, Weiss RG. Reduced myocardial creatine kinase flux in human myocardial infarction: an in vivo phosphorus magnetic resonance spectroscopy study. Circulation. 2009;119:1918–1924. doi: 10.1161/CIRCULATIONAHA.108.823187

37. Treibel Thomas A, Badiani S, Lloyd G, Moon James C. Multimodality Imaging Markers of Adverse Myocardial Remodeling in Aortic Stenosis. JACC: Cardiovascular Imaging. 2019;12:1532–1548. doi: 10.1016/j.jcmg.2019.02.034

38. Beyerbacht HP, Lamb HJ, van der Laarse A, Vliegen HW, Leujes F, Hazekamp MG, de Roos A, van der Wall EE. Aortic Valve Replacement in Patients with Aortic Valve Stenosis Improves Myocardial Metabolism and Diastolic Function. Radiology. 2001;219:637–643. doi: 10.1148/radiology.219.3.r01jn25637

39. Mahmod M, Francis JM, Pal N, Lewis A, Dass S, De Silva R, Petrou M, Sayeed R, Westaby S, Robson MD, et al. Myocardial perfusion and oxygenation are impaired during stress in severe aortic stenosis and correlate with impaired energetics and subclinical left ventricular dysfunction. J Cardiovasc Magn Reson. 2014;16:29. doi: 10.1186/1532-429x-16-29

40. Glaser N, Persson M, Franco-Cereceda A, Sartipy U. Cause of Death After Surgical Aortic Valve Replacement: SWEDEHEART Observational Study. Journal of the American Heart Association. 2021;10:e022627. doi: 10.1161/JAHA.121.022627

41. Glaser N, Persson M, Jackson V, Holzmann MJ, Franco-Cereceda A, Sartipy U. Loss in Life Expectancy After Surgical Aortic Valve Replacement: SWEDEHEART Study. Journal of the American College of Cardiology. 2019;74:26–33. doi: 10.1016/j.jacc.2019.04.053

42. Huded CP, Arnold SV, Chhatriwalla AK, Saxon JT, Kapadia S, Yu X, Webb JG, Thourani VH, Kodali SK, Smith CR, et al. Rehospitalization Events After Aortic Valve Replacement: Insights From the PARTNER Trial. Circulation: Cardiovascular Interventions. 2022;15:e012195. doi: 10.1161/CIRCINTERVENTIONS.122.012195

43. Lancellotti P, Magne J, Dulgheru R, Clavel M-A, Donal E, Vannan MA, Chambers J, Rosenhek R, Habib G, Lloyd G, et al. Outcomes of Patients With Asymptomatic Aortic Stenosis Followed Up in Heart Valve Clinics. JAMA Cardiology. 2018;3:1060–1068. doi: 10.1001/jamacardio.2018.3152

44. Banovic M, Putnik S, Penicka M, Doros G, Deja MA, Kockova R, Kotrc M, Glaveckaite S, Gasparovic H, Pavlovic N, et al. Aortic Valve Replacement Versus Conservative Treatment in Asymptomatic Severe Aortic Stenosis: The AVATAR Trial. Circulation. 2022;145:648–658. doi: 10.1161/circulationaha.121.057639

45. Kang D-H, Park S-J, Lee S-A, Lee S, Kim D-H, Kim H-K, Yun S-C, Hong G-R, Song J-M, Chung C-H, et al. Early Surgery or Conservative Care for Asymptomatic Aortic Stenosis. New England Journal of Medicine. 2019;382:111–119. doi: 10.1056/NEJMoa1912846

46. Hillis GS, McCann GP, Newby DE. Is Asymptomatic Severe Aortic Stenosis Still a Waiting Game? Circulation. 2022;145:874–876. doi: 10.1161/circulationaha.121.058598

47. Neubauer S, Horn M, Pabst T, Harre K, Strömer H, Bertsch G, Sandstede J, Ertl G, Hahn D, Kochsiek K. Cardiac high-energy phosphate metabolism in patients with aortic valve disease assessed by 31P-magnetic resonance spectroscopy. J Investig Med. 1997;45:453–462.

48. Edwards. The PROGRESS trial. www.edwards.com/healthcare-professionals/trial/progress-hcp. 2024. Accessed December.

